# Viral rebound among patients receiving COVID-19 convalescent plasma for treatment of Covid-19 in Uganda

**DOI:** 10.1101/2023.05.16.23290033

**Authors:** Patricia Alupo, Winters Muttamba, Levi Mugenyi, Ivan Kimuli, Katagira Winceslaus, Bruce Kirenga

## Abstract

**Background:** Viral rebound has been reported in people infected with COVID-19 treated with nirmatrelvir/ritonavir, and some cases been reported in patients who did not receive any antiviral treatment. Since the course of COVID-19 has not yet been well defined, we evaluated the incidence of viral rebound among COVID-19 patients treated with COVID-19 Convalescent Plasma (CCP) in Uganda.

**Methods:** In the CCP trail, 136 patients were enrolled between 21^st^ September 2020 and 2^nd^ December 2020 who presented to the Mulago National Referral COVID-19 treatment unit. Patients with a positive SARS-CoV-2 reverse transcriptase (RT)-PCR test irrespective of disease severity were hospitalised and randomised to receive either COVID-19 CCP plus standard of care (SOC) or SOC alone. SARS-CoV-2 RT-PCR was done at baseline and on days 3, 5, 7, 14 and 28 post randomisation or until two consecutive negative RT-PCR results were obtained, whichever occurred first. We analysed for occurrence of viral rebound. Viral rebound was defined as a positive SARS-CoV-2 RT-PCR test following a prior negative test.

**Findings:** 20% of the participants had viral rebound. Viral rebounders were predominantly male. The median age was 45-64 years and they had at least one co-morbidity. There was no difference in the rebound rates in the study arms, and participants with hypertension had more rebound rates compared to those with other co-morbidities.

**Interpretation:** Viral RNA rebound was common among patients receiving CCP. Viral rebound may be a result of the biphasic nature of COVID-19 infection, and not a consequence of the therapeutic interventions.

## Introduction

There have been documented reports of COVID-19 rebound in patients receiving Paxlovid, and subsequent alluding to the fact that COVID-19 rebound is not related to Paxlovid treatment. (1)(2)(3).However, this rebound phenomenon has not satisfactorily been understood, and has thus far not been evaluated in patients receiving COVID-19 Convalescent plasma.

We would thus like to share our perspective on COVID-19 viral rebound from Uganda, where we had patients receiving non-conventional COVID-19 antiviral therapy; namely COVID-19 convalescent plasma in a clinical trial setting. Since the declaration of COVID-19 as a global pandemic (4), several reports have been made concerning the clinical features, treatment options and outcomes of patients with COVID -19 pneumonia (4)(5).

However thus far, the natural course of viral and symptom trajectories of COVID-19 infection is not well understood. From the outset, SARS -CoV-2 was expected to have a monophasic course, and to immunize the host at least transiently, following an infection with the virus, as was described concerning other coronaviruses (6) (7). Following the first cases of SARS-CoV-2 reactivation that were reported in South Korea in 2020, there has been renewed interest in understanding of the disease trajectory (8). Since then, more cases have since been reported with patients who were previously declared COVID-19 negative having a rebound of symptoms and viral RNA load (3), and efforts been made to characterize them (9) .Of interest, viral rebound has been reported among patients receiving Nimatrelvir /Ritonavir (Paxlovid), with some reports showing patients getting worse after treatment completion (9), and some gone ahead to demonstrate presence of viral and symptom rebound in the absence of antiviral treatment (3). Amidst limited therapeutic options for COVID-19, the COVID-19 research group based at Makerere University Lung Institute explored cheaper and easily accessible therapeutic options that could lead to earlier viral clearance, and patient recovery. A randomized open-label clinical trial was conducted to determine if administration of CCP to patients who were reverse transcriptase (RT)-PCR-positive at hospitalization would lead to earlier clearance of SARS-Cov-2 and better clinical outcomes. We evaluated the rate of viral rebound among the patients who were enrolled on this trial, to evaluate the incidence of viral rebound in this patient population.

## Methods

The CCP trial was an open-label, randomised clinical trial conducted at Mulago National Referral Hospital (MNRH) COVID-19 Treatment Unit. The trial included patients with documented SARS-CoV-2-positive RT-PCR performed at the trial laboratory of Makerere University Department of Immunology and Molecular Biology. We excluded patients with a prior diagnosis of IgA deficiency and those unable to participate in follow-up procedures. Permuted block randomisation with varying sizes of blocks was used to randomly assign eligible participants to receive either CCP plus standard of care (CCP+SOC) or standard of care only (SOC). All participants provided written informed consent prior to enrolment.

### Intervention

Participants were enrolled between 21^st^ September 2020 and 2^nd^ December 2020, and the trial enrolled a total of 136 participants (69 received intervention CCP and standard of care, 67 received only standard of care), and followed them up for a period of 28 days. Patients with a positive SARS-CoV-2 reverse transcriptase (RT)-PCR test irrespective of disease severity were hospitalised and randomised to receive either COVID-19 CCP plus standard of care (SOC) or SOC alone. SARS-CoV-2 RT-PCR was done at baseline and on days 3, 5, 7, 14 and 28 post randomisation or until two consecutive negative RT-PCR results were obtained, whichever occurred first. We analysed for occurrence of viral rebound. Viral rebound was defined as a positive SARS-CoV-2 RT-PCR test following a prior negative test. More detailed study procedures are described in the publication of the primary study(10).

### Sample size calculation

The sample size was calculated based on the primary outcome of the CCP trial (10). The primary outcome of the randomised clinical trial was viral clearance. The trial was planned to detect a minimum hazard reduction in the primary outcome of 40% in the CCP (intervention) arm, equivalent to an HR of 0.6. With a power of 80%, a two-tailed type 1 error (alpha) of 0.05, a patient accrual period of 3 months, a total study time of 6 months, a ratio of accrual to total time of 0.5, with no anticipated cross-over (no dropout of, or drop-in, the intervention arm), a ratio of n1 to n2 of 1:1, and equal enrolment rate in both arms, the required unadjusted sample size per group/arm was estimated to be 66 patients, giving a total of 132. Given that the trial was testing a potential therapy for a disease with no proven therapy and that all participants will be inpatients, a minimal loss-to-follow-up of 3% is anticipated, and after adjusting for it the total number of patients to be enrolled and randomised totalled to 136 (68 per arm). More details concerning the randomization process can be obtained from the primary study publication (10).

### Statistical Analysis

Proportions were used to summarize both demographic and clinical characteristics of the study participants. The study outcome was the number of COVID-19 rebounds recorded over the follow-up period. Time to each rebound was obtained and presented using median (interquartile range, IQR). Also, the number and proportion of participants with at least one rebound was presented by sex, age groups, commodities, and oxygen saturation. Poisson regression using person time in years as an offset variable was used to estimate incidence rate and incidence rate ratios with 95% confidence intervals (CI). Incidence of rebound was presented overall and by participants characteristics. STATA version 15 was used for analysis.

## Results

We enrolled 136 participants in the trial, and 71 % of them were male. The larger part of the participants was in the age category of 45-64 years. 58% of the participants had at least one comorbidity. Hypertension was the most prevalent comorbidity (36.1%), followed by diabetes mellitus (23.5%) and HIV (11.0%) respectively. Regarding management approaches, 48.5% of the participants needed supplemental oxygen therapy, (Table1)

**Table 1.** Table showing the number and time to COVID 19 rebound by sex, age group, co-morbidities, and disease severity as demonstrated by need for oxygen therapy in this patient population.

|  | **Number of COVID Patients**<br>**n=136 (%)** | **Had at least one rebound**<br>**n (%)** | **Total number of rebound cases** | **Time (days) to rebound**<br>**median (IQR)** |
| --- | --- | --- | --- | --- |
| **Sex** |  |  |  |  |
| Male | 97 (71.3) | 21 (21.7) | 22 | 2 (2, 2) |
| Female | 39 (28.7) | 7 (18.0) | 7 | 2 (2, 2) |
| **Age Categories (years)** |  |  |  |  |
| <25 | 5 (3.7) | 0 (0.0) | 0 | --- |
| 25-44 | 40 (29.4) | 8 (20.0) | 9 | 2 (1.5, 2) |
| 45-64 | 61 (44.9) | 15 (24.6) | 15 | 2 (2, 2) |
| 65+ | 30 (22.1) | 5 (16.7) | 5 | 2 (2, 2) |
| **<u>Comorbidities:</u>** |  |  |  |  |
| At least one comorbidity | 79 (58.1) | 18 (22.8) | 18 | 2 (2, 2) |
| Hypertension | 49 (36.0) | 11 (22.5) | 11 | 2 (2, 7) |
| Diabetes | 32 (23.5) | 5 (15.6) | 5 | 2 (2, 2) |
| Asthma | 5 (3.7) | 1 (20.0) | 1 | 2 (2, 2) |
| HIV | 15 (11.0) | 4 (26.7) | 4 | 2 (2, 2) |
| Tuberculosis | 4 (2.9) | 0 (0.0) | --- | --- |
| Cancer | 1 (0.7) | 0 (0.0) | --- | --- |
| COPD | 0 (0.0) | --- | --- | --- |
| Sickle cell | 0 (0.0) | --- | --- | --- |
| Kidney disease | 0 (0.0) | --- | --- | --- |
| **<u>Oxygen therapy</u>** |  |  |  |  |
| Zero oxygen | 70 (51.5) | 14 (20.0) | 15 | 2 (2, 2) |
| Oxygen 1-5 L/min | 35 (25.7) | 9 (25.7) | 9 | 2 (2, 2) |
| Oxygen 6-10 L/min | 8 (5.9) | 0 (0.0) | 0 | --- |
| Oxygen 11-15 L/min | 20 (14.7) | 4 (20.0) | 4 | 2 (2, 4.5) |
| Oxygen >15 L/min | 3 (2.2) | 1 (33.3) | 1 | 2 (2, 2) |

We observed that 20% of the participants had viral rebound, with higher rebound rates observed in males, participants aged between 45-64 years of age, and those who had at least one co-morbidity. There was no difference in rebound rates between the study arms. Among those with co-morbidities, participants with HIV had the highest rebound rates (26.1%), followed by hypertension (22.5%), and asthma 20.0% (Table 2)

**Table 2:** Incidence of Viral rebound, overall and by study arm and baseline characteristics.

|  | **Number of rebound cases** | **Total person years** | **Incidence of rebound per person year (95% CI)** | **Incidence Rate Ratio (IRR) (95% CI)** | **P value** |
| --- | --- | --- | --- | --- | --- |
| **Overall** | **29** | **4.99** | **5.81 (4.03 – 8.36)** |  |  |
| **Arm** |  |  |  |  |  |
| Control | 14 | 2.41 | 5.82 (3.45, 9.83) | 1.00 |  |
| Intervention | 15 | 2.59 | 5.79 (3.49, 9.61) | 1.00 (0.48, 2.06) | 0.990 |
| **Sex** |  |  |  |  |  |
| Male | 22 | 3.53 | 6.24 (4.11, 9.48) | 1.00 |  |
| Female | 7 | 1.47 | 4.77 (2.27, 10.0) | 0.76 (0.33, 1.79) | 0.535 |
| **Age group (years)** |  |  |  |  |  |
| <25 | 0 | 0.27 | --- |  |  |
| 25-44 | 9 | 1.64 | 5.49 (2.86, 10.56) | 1.00 |  |
| 45-64 | 15 | 2.07 | 7.26 (4.38, 12.04) | 1.32 (0.58, 3.02) | 0.508 |
| 65+ | 5 | 1.02 | 4.92 (2.05, 11.82) | 0.90 (0.30, 2.67) | 0.843 |
| **Comorbidities:** |  |  |  |  |  |
| At least one comorbidity | 18 | 2.98 | 6.03 (3.80, 9.58) | 1.10 (0.52, 2.34) | 0.798 |
| Hypertension | 11 | 2.11 | 5.20 (2.88, 9.39) | 0.83 (0.39, 1.76) | 0.631 |
| Diabetes | 5 | 0.78 | 6.45 (2.68, 15.49) | 1.13 (0.43, 2.97) | 0.799 |
| Asthma | 1 | 0.18 | 5.45 (0.77, 38.67) | 0.94 (0.13, 6.88) | 0.948 |
| HIV | 4 | 0.48 | 8.34 (3.13, 22.23) | 1.51 (0.52, 4.33) | 0.447 |
| **Oxygen therapy** |  |  |  |  |  |
| Zero oxygen | 15 | 2.65 | 5.65 (3.41, 9.37) | 1.00 |  |
| Oxygen 1-5 L/min | 9 | 1.57 | 5.72 (2.98, 11.0) | 1.01 (0.44, 2.31) | 0.976 |
| Oxygen 6-10 L/min | 0 | 0.10 | --- | --- | --- |
| Oxygen 11-15 L/min | 4 | 0.60 | 6.79 (2.55, 18.09) | 1.20 (0.40, 3.62) | 0.744 |
| Oxygen >15 L/min | 1 | 0.08 | 13.04 (1.84, 92.54) | 2.31 (0.30, 17.47) | 0.418 |

Interestingly, when categorized based on disease severity as assessed by the need for supplemental oxygen therapy, we observed a similar rebound percentage among both those that did and did not receive oxygen supplementation as part of their management. The median time to rebound across both patient arms was 2 days.

## 5. Discussion

In this study evaluating Covid 19 convalescent plasma as a therapeutic approach to the management of COVID-19, 69 participants received CCP and standard of care, and 67 received only standard of care. We found that viral rebound was relatively common with 20% of the participants experiencing COVID-19 viral rebound. The median time to rebound was 2 days. Rebound cases were more prevalent among patients who had at least one co-morbidity, with HIV co-infected patients having the highest rebound rates.

The percentage of rebound was much higher than registered in the studies cited herein, ranging from 2.81-12% (11)(3,12)(3). Given both trials were conducted in 2020 where the predominant COVID-19 variant was alpha, the higher viral rebound rate in this population compared to the that of say Rinki *et al* could be as a result of the difference in disease severity of the participants involved. The CCP trial enrolled participants across the entire disease spectrum from mild to severe disease compared to those enrolled in the study by Rinki and colleagues who only had mild to moderate disease.

Possible etiologies that could elucidate the viral relapse include the potential for the virus lodging in varied anatomic spaces and subsequently the viral shedding happening slowly. It could be as result of re-infection with the virus(13)(14), and the possibility of it being as a result of resistance to the CCP cannot be ruled out.

This study had limitations, the first being the small sample size, and the second being that it was conducted during the alpha wave, and as such the subsequent variants were not catered for. The third limitation is that that we did not capture symptom rebound, and thus could not link the viral rebound to symptom rebound in this cohort. This therefore highlight the need for larger, more robust studies that include all variants to evaluate both viral and symptom relapse, and the pathophysiology at play in this with the co-morbidities, especially HIV.

## Conclusion

Our findings show that COVID 19 viral rebound is relatively common among patients receiving COVID 19 Convalescent Plasma. Viral load rebound is likely an exhibition of the biphasic nature of some variants of COVID 19. Further research is needed to better understand the trajectory of COVID-19 infection.

## Data Availability

All data produced in the present study are available upon reasonable request to the authors

## Acknowledgments

The funders of the study-Makerere University Research and Innovations fund, and the patients that accepted to participate in the study.

